# Research opportunities in precision oncology: Perspectives on biospecimen and genomic data sharing from adults with cancer in Ireland

**DOI:** 10.1101/2024.11.27.24318092

**Authors:** Shane O’Grady, Jessica C. Ralston, Eadaoin McKiernan, Frances J. Drummond, Jan Rynne, Derick Mitchell, William M. Gallagher, Amanda Drury, Walter Kolch

**Affiliations:** Systems Biology Ireland, School of Medicine, University College Dublin, Dublin, Ireland; Breakthrough Cancer Research, Cork, Ireland; Chronic Lymphocytic Leukaemia (CLL) Ireland, Dublin, Ireland; The Irish Platform for Patient Organisations, Science and Industry (IPPOSI), Dublin, Ireland; UCD School of Biomolecular and Biomedical Science, UCD Conway Institute, University College Dublin, Dublin, Ireland; School of Nursing, Psychotherapy and Community Health, Dublin City University, Dublin, Ireland; Conway Institute of Biomolecular and Biomedical Research, University College Dublin, Dublin, Ireland

## Abstract

Although surveys of people living with cancer in other nations have generally found a high degree of willingness to donate to research services such as genomic databases and biobanks, these results can vary from country to country. Additional questions also remain surrounding issues such as preferred model of consent, reporting of findings and concerns surrounding potential commercial application of health data. To address some of these gaps in our current knowledge, we collected responses from 176 people living with and beyond cancer in Ireland over a 16-month period. Twenty-eight percent of our survey respondents had previously donated a biological sample to research, with the majority indicating that they did so for altruistic reasons. We found that the vast majority of those who had not previously donated samples would do so if given the opportunity, suggesting that there is a significant untapped pool of potential sample donors, and increased efforts by researchers and clinicians to recruit these individuals, in an ethically acceptable way, could yield a substantial improvement in the availability of biological samples and data for research. There was also a strong preference among respondents for total transparency with personal health data, with the vast majority wanting to know of any risk factors identified in their genome, even if these risks were not medically actionable. A strong level of trust in both the clinical and scientific community was also observed in the responses, with most indicating that this played a major role in influencing their decision to donate. Finally, we found that although most respondents did not have issues with the involvement of a commercial entity in the donation process, there was still a sizeable percentage (26%) who did have some reservations.

## Introduction

Genomic analysis is by now a well-established cornerstone of modern cancer research efforts, greatly enhancing our understanding of tumour biology and assisting in the development of new treatment options (1). In addition, increased sequencing availability has also led to a rapid expansion in its clinical adoption (2) and many people diagnosed with cancer now undergo some level of analysis as part of their diagnostic workup, ranging from relatively simple analysis of a small gene-panel all the way up to whole-exome or even whole-genome sequencing (3, 4).

The continued development and implementation of genomic analysis in both bench and bedside settings is only possible with the availability of sufficiently sized cohorts of biological data and samples. Whether these are derived from direct recruitment of participants to a specific study or from a more generalised approach such as tissue biobanks, it is ultimately dependent on the willingness of individuals to engage with clinicians and researchers and donate their biological samples and personal health data. Studies conducted in other countries have generally found people with cancer to be highly willing to engage with researchers and donate biological samples and data for research purposes (5–7), often citing altruistic motivating factors such as a desire to improve treatment options for other people with cancer (8, 9). However, while willingness to donate is generally found to be high in these studies, it is not universal and significant variation exists between countries. As an example, results from the 2010 Eurobarometer study showed that willingness to provide information to a biobank varied dramatically between countries, ranging from 92% in Iceland to 24% in Turkey, with an EU-wide average rate of 46% (10).

Previous reports have highlighted concerns around data privacy and fears that personal health or genetic data could be misused in some way (e.g. resulting in discrimination by insurance or loan companies) as potential factors that may influence an individual’s decision to donate (5, 11, 12). The involvement of a commercial entity in sample collection and storage, especially when concerning genetic data, may also cause some trepidation (13).

The implementation of genomic analysis to the clinic in Ireland has not been without difficulties, with some arguing that Ireland is somewhat behind the curve in several key areas (14), while a report commissioned by the Irish Cancer Society has also highlighted major issues with access to genetics services (15). Additionally, some aspects of key research infrastructure are also lacking. This was highlighted in a recent report on the current biobanking landscape in Ireland, which emphasised significant challenges currently hampering biobanking efforts in the country, including a lack of primary biobanking legislation and national biobank office as well as issues with long-term funding and staff retention (16).

More encouragingly, previous reports do suggest a strong degree of willingness amongst the public to engage with researchers and provide biological samples and personal health data. For instance, work from the Irish Platform for Patients’ Organisations, Science & Industry (IPPOSI) has indicated that the public are generally willing to allow sharing of their health-data for research purposes, provided there is a public benefit and stringent protections are provided (17).

Addressing issues within the genomics and biobanking landscapes will require up-to-date information on the views of people living with cancer on topics such as participation in research, preferred model of consent and disclosure of findings. The aim of this study was to address this knowledge gap and to understand the behaviour and factors influencing the donation of medical samples and data to research among people living with and beyond cancer in Ireland.

## Methods

### Study design

The survey was designed to collect data from adult participants living in Ireland, with a lived experience of cancer. Participants were designated as “Previous donors” or “Non donors” based on their response to a question on whether they had previously donated a sample to research. The survey was designed with a branching structure, allowing for segregation of data based on previous donation status.

### Participants and setting

A total of 176 survey participants were recruited through mailing lists of Irish cancer charities as well as via social media channels. Respondents were asked to self-select as a “Cancer patient” or “Cancer survivor in remission”. Survey data was gathered over a 16-month period between September 2022 and December 2023.

### Data collection

Interested participants were directed to an online platform (Google Forms), which was used to collect their responses anonymously. The survey home page provided prospective participants with a description of the study aims, overview of questions to be asked and statements on survey data-protection. Participants were required to tick a box to indicate consent before proceeding with the survey. Only fully completed responses were collected.

Data is displayed for the full cohort for initial demographics questions (Fig. 1), as well as data on trust levels and opinions on biobanking, consent & commercialisation (Fig. 4 & Fig. 5), while data regarding the donation process is displayed separately for these two groups (Fig. 2 & Fig. 3).

**Figure 1.**
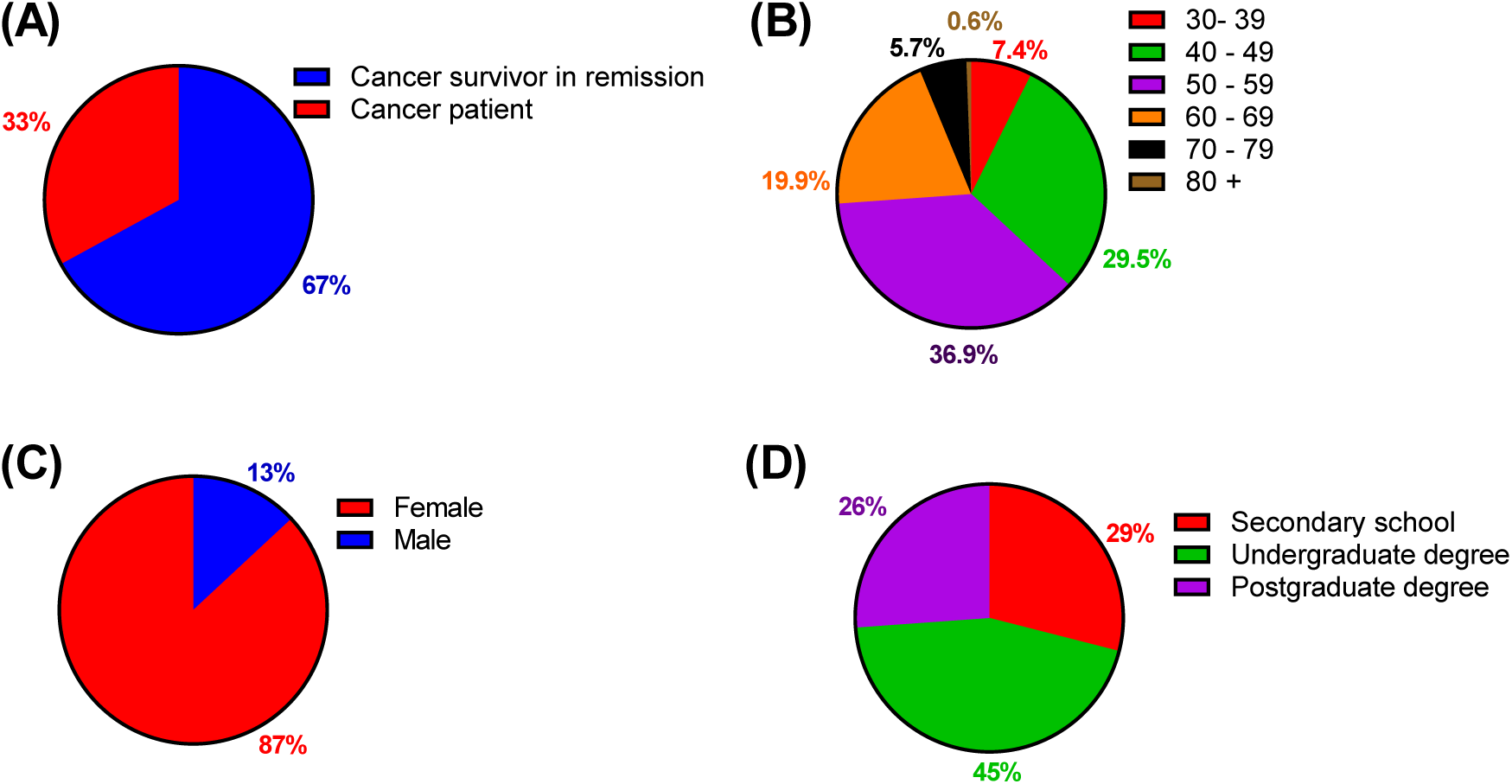
Respondent demographics. Cancer status (A), age-brackets (B), gender identity (C) and highest educational attainment (D) of survey respondents (N=176).

**Figure 2.**
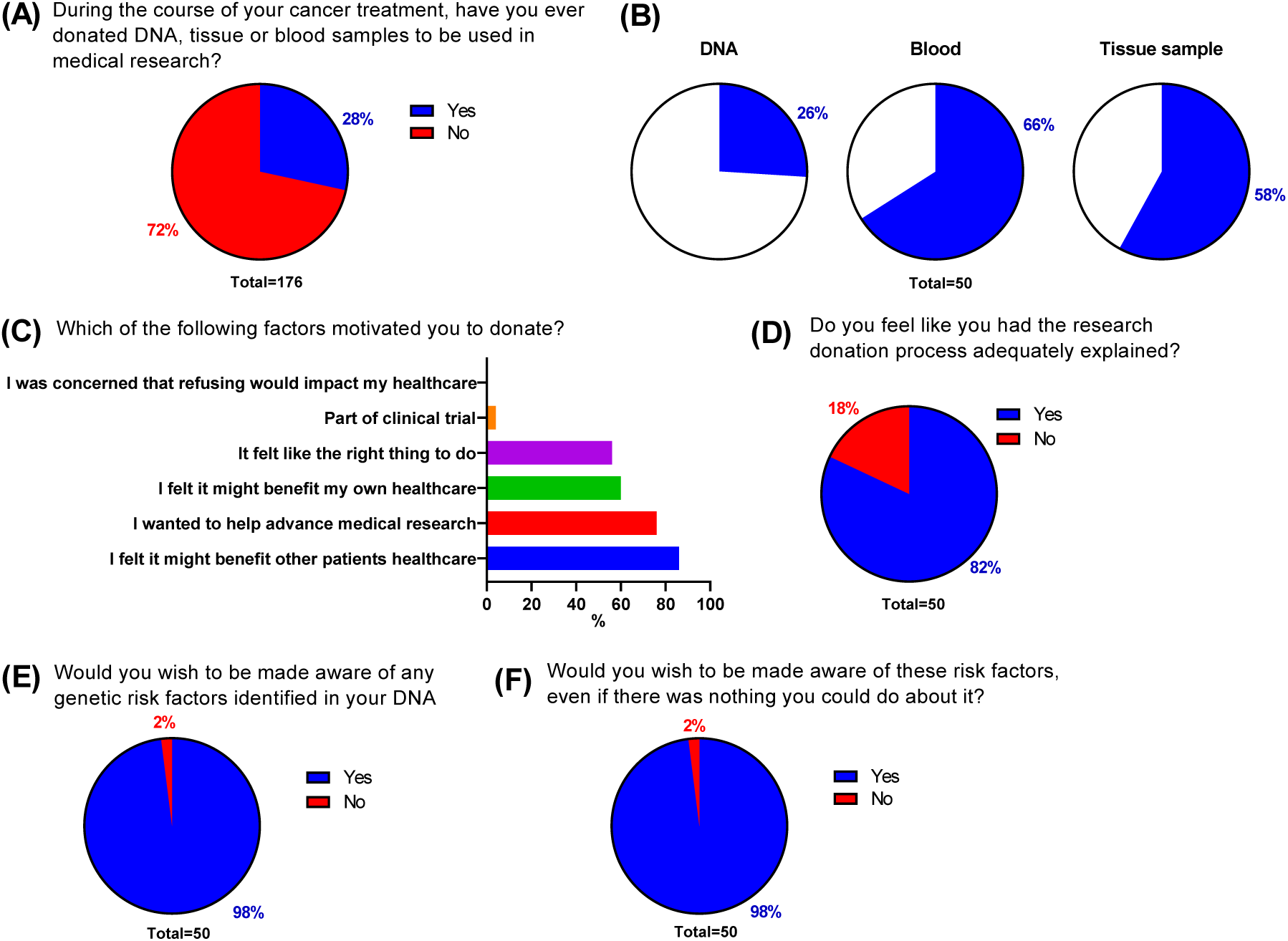
Previous donors. Survey cohort was subdivided based on whether they had previously donated samples or data to research (A). Previous donors (N=50) were surveyed on sample type (B), motivations for donating (C), satisfaction regarding explanations of donation process (D), and desires to be made aware of genetic findings (E) and non-actionable genetic findings (F).

**Figure 3.**
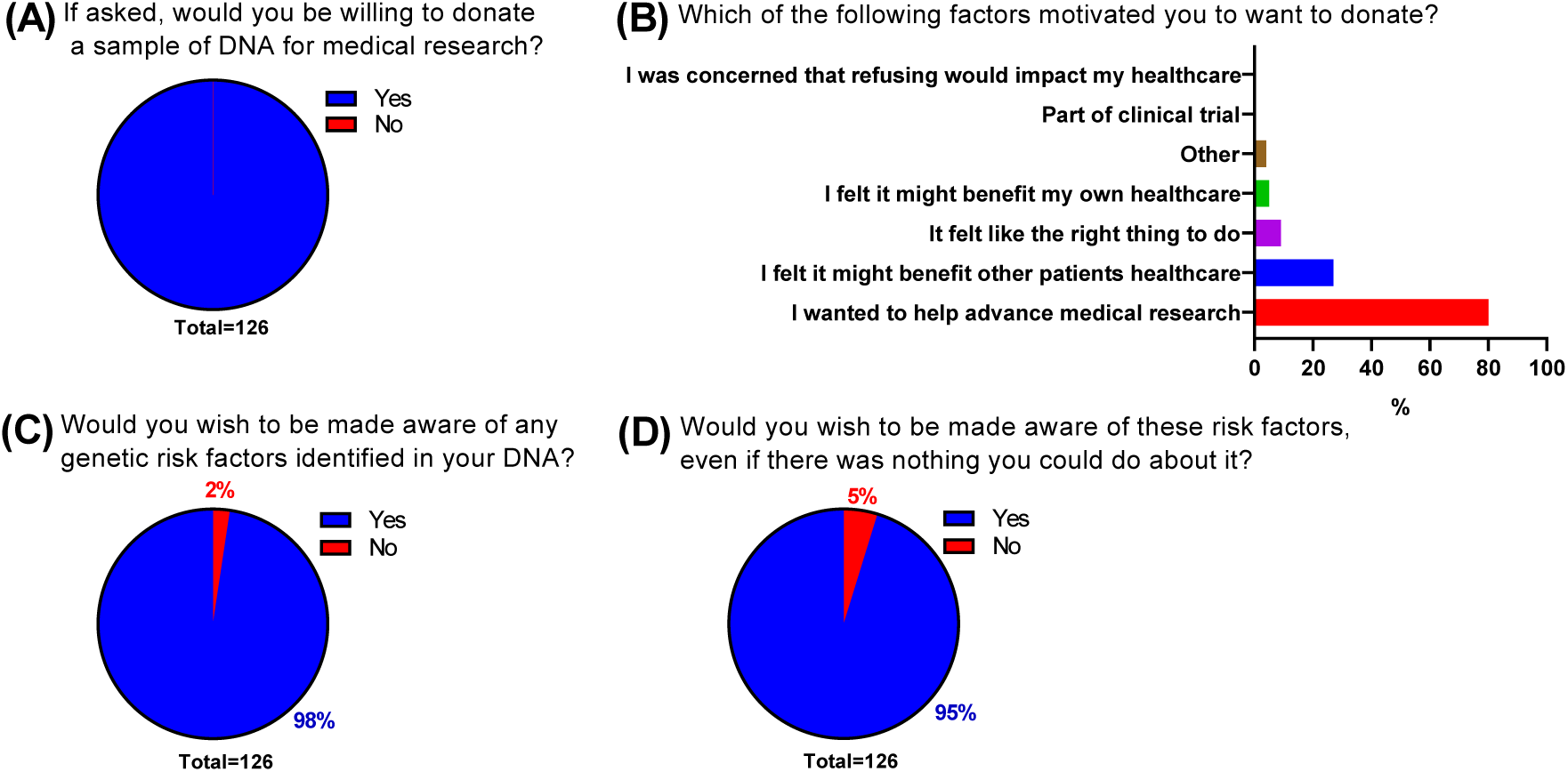
Non-donors. Survey respondents who had not previously donated samples or data to research (N=126) were polled on if they would do so if given the chance (A), factors that would motivate them to donate (B), and desires to be made of aware of genetic findings (C) and non-actionable genetic findings (D).

**Figure 4.**
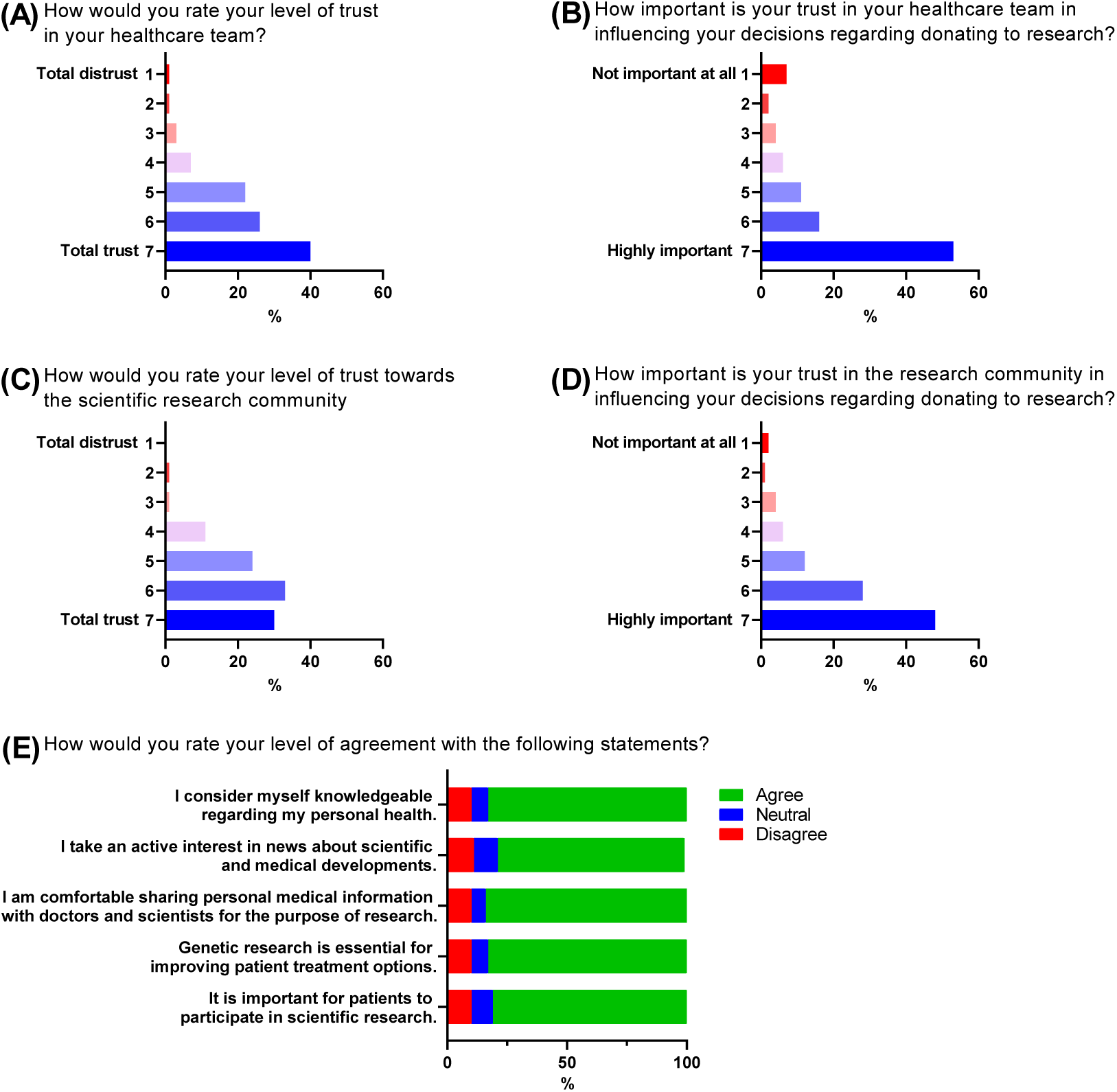
Trust in healthcare teams and researchers. Survey respondents (N=176) were asked about on their level of trust in their healthcare team (A) and scientific research community (C) as well as the influence of these trust levels on decision making (B,D). Interest and opinions regarding research & personal health was also assessed with a series of 5 statements (E).

**Figure 5.**
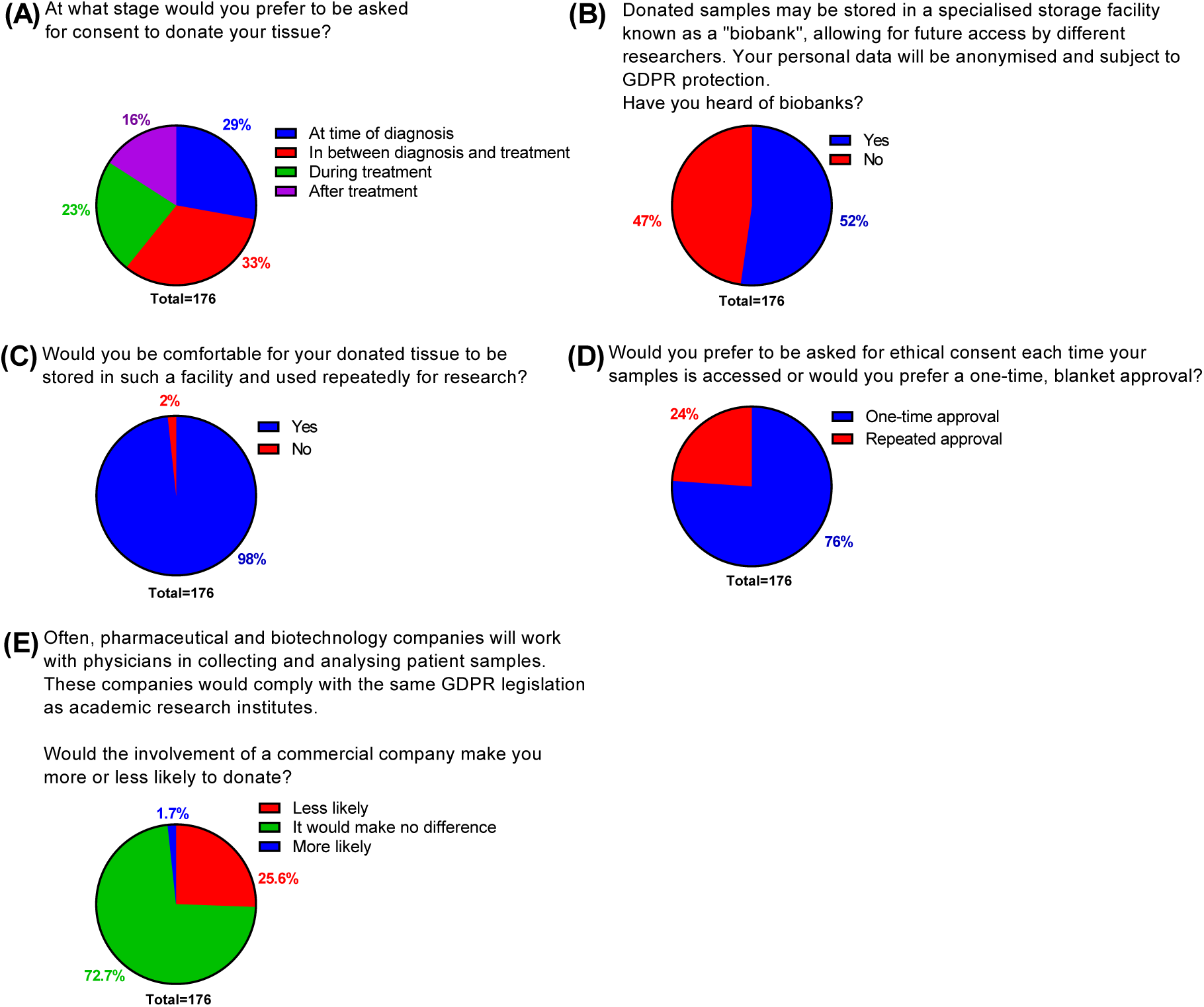
Consent, biobanking & commercialisation. Survey respondents (N=176) were polled on their preferred time to grant consent (A), whether they had heard of biobanks (B), willingness to donate to biobanks (C), preference on consent approval (D) and opinions on commercial involvement in the donation process (E).

The majority of data was collected as single-answer questions except questions regarding type of sample donation and motivating factors to donate, which allowed for multiple selections. Data regarding levels of trust in healthcare and research teams was gathered using a 7-point Likert scales, ranging from 1 indicating “Total distrust” and 7 indicating “Total trust”. Data regarding participant opinions on research & personal health was initially collected using a 5-point scale ranging from “Strongly disagree” to “Strongly agree”. Overall agreement levels were generated by combining figures for “Strongly disagree” with “Disagree” and “Strongly agree” with “Agree”, to create a 3-point scale of “Disagree”, “Neutral” and “Agree”

### Data analysis

Data was analysed using SPSS 29. Data was treated as either nominal (gender identity, previous donation status, cancer status, opinions on consent, biobanking, return of findings and commercialisation) or ordinal (educational attainment, age, trust levels, agree/disagree statements) categorical data types. Descriptive statistics are presented as percentages. Denominators varied as some questions were only applicable for certain group.

Relationships between variables was tested using Spearman correlation (when testing two ordinal variables) or Chi-square (when testing mixed nominal and ordinal variables). Significance was determined as p < 0.05.

Graphs were created using GraphPad Prism 8.

## Ethics approval

This study received “low-risk” ethics approval from University College Dublin’s Human Research Ethics Committee – Sciences (HREC-LS).

## Results

### Overall demographics

In total, 176 individual responses were collected. Respondents were primarily individuals whose cancer was in remission (67%, Fig. 1A). Our survey captured responses from individuals across a wide range of age categories; (Fig. 2B), with the highest response rate received from under 50 – 59 (36.9%) followed by 40 – 49 (29.5%), 60 – 69 (19.9%), 70 – 79 (5.7%), 30 – 39 (7.4%) and finally 80+ (0.6%). Our cohort was skewed towards female, with 87% of respondents identifying as female (Fig. 1C) and only 13% identifying as male. The pattern of low response rate from males remained despite outreach efforts to support groups catering to prostate and other forms of male-associated cancer. Finally, our cohort had high levels of third-level education, with 45% indicating an undergraduate degree as their highest level of educational attainment and 26% progressing to postgraduate studies (Fig. 1D). The remaining 29% achieved a secondary school qualification as their highest qualification. The most recent available Census data indicates 48% of the Irish population possesses a third-level degree, while 27% reporting upper secondary as their highest educational level achieved (18).

### Previous donors

When asked if they had ever donated DNA, tissue or blood samples to be used in medical research, 28% of respondents indicated that they had previously donated a sample (Fig. 2A). Within this subgroup who had donated, the most common donation type was a blood sample (66%, Fig. 2B) followed by tissue sample (58%) and DNA (26%). The most common reason cited for choosing to donate was “I felt it might benefit other patients’ healthcare” (86%, Fig. 2C), followed by “I wanted to help advance medical research” (76%), “I felt it might benefit my own healthcare” (60%) and “It felt like the right thing to do” (56%). A small number (4%) also indicated that their donation had been part of a clinical trial. Of those who had previously donated, the majority (82%, Fig. 2D) felt that the donation process was adequately explained to them. There was, however, a sizable percentage (18%) who did not agree and wished for a better explanation process prior to donation.

Finally, the vast majority (98%, Fig. 2E) of those who had donated, wished to be made aware of any genetic risk factors found within their DNA. Interestingly, this did not change when respondents were asked about being made aware of non-actionable genetic risks (Fig. 2F).

### Non-donors

Amongst the 72% of respondents who indicated that they had never donated samples to research, there was universal agreement (100%, Fig. 3A) that they would be willing to donate a sample if given the chance, suggesting a strong desire in the cohort to participate in research.

When polled on what factors would influence them to donate, there were some differences in comparison to those who had already donated. By far the most common reason was “I wanted to help advance medical research” (80%, Fig. 3B), with much lower percentages answering for each of the remaining options. This contrasted with the subgroup who had donated, where their answers were more evenly distributed between the options (Fig. 2C).

### Trust

To get a better sense of our cohort’s views on the healthcare and scientific communities, we next asked responders about their level of trust in these two groups and what impact that trust would have on their choices regarding sample donation. Trust levels in the healthcare community were high, with 40% answering with the maximum score of 7 and another 26% answering with a 6 (Fig. 4A). When asked to rank how important this level of trust was in influencing the decision to donate, with 1 indicating “Not important at all” and 7 indicating “Highly important”, 53% and 16% answered with a 7 or 6 respectively (Fig. 4B).

When these questions were asked about the scientific community, there was a lower percentage of respondents indicating “Total trust” (30%, Fig 4C) in comparison to the healthcare community (Fig. 4A). This was balanced out by higher percentage of people answering with a 6 (30%), meaning that approximately 2/3 of respondents had a very high level of trust in both the healthcare and scientific communities, albeit with minor differences in how these scores were distributed. The impact of trust level in the scientific community when it comes to influencing an individual’s decision to donate was also observed to be important, with 48% and 28% answering with a 7 or 6, respectively (Fig. 4D). We did not observe any significant relationship between trust levels and participant age, gender identity or educational attainment.

We also asked respondents a series of opinion questions to assess their level of interest in science and viewpoints on research. Responses were highly positive (Fig. 4E), with the level of agreement with each of the 5 statements ranging between 78% (“I take an active interest in news about scientific and medical developments”) and 85% (“I am comfortable sharing personal medical information with doctors and scientists for the purpose of research”).

We observed significant correlations between educational level and agreement with 3 out of 5 statements regarding viewpoints on research, with higher levels of education correlating with increased agreement with the statements “I consider myself knowledgeable regarding my personal health”, “I take an active interest in news about scientific and medical developments” and “It is important for patients to participate in scientific research” (Fig. S1).

### Biobanking, consent & commercialisation

We also queried respondents on their opinions regarding consent timing and the usage of biobanks. We first asked at what stage of their diagnosis and treatment would they prefer to be asked for consent to donate tissue. Overall, more than 1 in 3 respondents 33% would prefer to be asked for consent to donate a sample “In between diagnosis and treatment” (Fig. 5A). This was followed by “At time of diagnosis” (29%), “During treatment” (23%) and “After treatment” (16%).

When we asked respondents if they had heard of tissue biobanks, just over half of respondents (52%, Fig. 5B) were aware of biobanks. The vast majority of respondents indicated that they would be comfortable for their tissue to be stored in a biobank and made available to researchers (98%, Fig. 5C). The majority of respondents preferred a one-time approval to allow researchers blanket access to their samples (76%, Fig. 5D). However, just under a quarter (24%) would prefer to be repeatedly asked for consent.

Finally, we asked if the involvement of commercial entities in the donation process would affect people’s willingness to donate. In our cohort, we found that although most people were not concerned with commercial involvement, stating that involvement of a commercial company would make no difference in their decision to donate (72.7%, Fig. 5E), approximately a quarter (25.6%) reported that commercial involvement would decrease their likelihood of donating.

## Discussion

Modern medicine is increasingly data-driven and there is perhaps no clearer example of that than in oncology, where large omics databases are crucial tools in developing the next generation of personalised treatment options and high-sensitivity diagnostic assays (19). The availability of such datasets would not be possible without the continued participation of people with a cancer diagnosis in altruistic sample donation, necessitating a clear understanding of not only their willingness to donate but also the motivating and deterring factors underlying this decision.

Results from this study demonstrate that people living with and beyond cancer in Ireland have extremely high levels of willingness to donate samples to research. We found that 100% of those surveyed would be willing to provide samples to research and only 2% would not be willing to donate to a tissue biobank. This enthusiastic willingness has also been observed not only in other countries (5, 7, 20, 21) but also in Ireland. A previous study of people attending cancer clinics in Ireland found that 79% would be willing to donate personal data or a blood sample as part of a clinical trial (22). Similarly, a survey across 4 Irish cancer centres found a majority of respondents expressed strong support for biomedical and genetic research, with 16% having previously donated a sample to research (23). More recently, a survey on people attending an Irish pigmented lesion clinic found that the majority of those with a melanoma diagnosis were willing to donate tissue and blood samples, even if it required an additional skin biopsy (24).

Comparing this to data from the general public, a cross-sectional telephone survey of the Irish populace found that 86% of respondents would agree to donate excess surgical tissue following a procedure (25) while a report from Cancer Trials Ireland in 2022 found that 80% of respondents would be willing to donate blood for clinical research (26). Encouragingly, this represented a 4% increase compared to data gathered by the same group just two years prior (26).

Despite this willingness, just 28% of respondents to our survey said that they had previously donated to research, indicating that there remains a great untapped pool of willing donors and that efforts to increase the rate of participation in research initiatives would greatly expand the availability of vital research data and materials.

Studies in other countries have attempted to quantify which attributes influence an individual’s willingness to donate tissue samples or data to research, often with mixed results. Whilst educational levels have been shown in many studies to be predictive of willingness to donate samples to research (5, 25, 27, 28), this is not universally observed (29–31). Results from this study would also suggest a link between educational attainment and research participation. Although we could not make a direct comparison between educational levels and willingness to donate to research (due to the lack of respondents indicating they would not donate), we did observe a significant correlation between educational attainment and agreeing that it was important to participate in scientific research.

Many studies have also highlighted a pattern of greater awareness and familiarity with relevant scientific concepts to be a significant factor in predicting willingness to participate in research. Individuals with greater familiarity of genetic concepts are more willing to donate genomic data to research (32, 33) and undergo genetic testing (34, 35), while studies of biobanking participation have often found levels of awareness of biobanking activity to significantly influence willingness to donate to a biobank (36). This is concerning in light of the fact that a recent survey of people living with cancer in Ireland found that the majority declared “little or no knowledge” of genetic testing (37), while a 2010 EU-wide survey found Ireland had slightly lower awareness of biobanks (31%) compared to the EU average of 34 % (10).

This suggests a role for a significant expansion in public educational efforts to increase the general awareness of the benefits of biobanks and enhance public understanding key concepts in genomics research. The importance of this goal was recognised in the HSE’s recently published “National Strategy for Accelerating Genetic and Genomic Medicine in Ireland”, which amongst many other recommendations designed to hasten the integration of genomics and personalised medicine into the Irish healthcare system, included a strategy on improving public awareness of genetics and genomics (38). In a similar manner, results from a “citizen’s jury” on the topics of

genomics, coordinated by Irish Platform for Patients’ Organisations, Science & Industry (IPPOSI), highlighted the importance of public awareness in all aspects of genomics in health care and health research (39). One potential avenue would be to improve availability of research infrastructure within hospitals and expand opportunities for interaction between patients and research staff, such as biobanking managers. A recent report highlighted the need to further develop Ireland’s biobanking infrastructure, providing a number of recommendations including the establishment of a national biobank office, delivering training programmes in biobanking competencies and providing meaningful opportunities for patient and public involvement (16).

The same report also touched on the issue of consent, including support for a broad-consent model for biobanking. We observed strong approval amongst our respondents for a one-time approval model of consent versus requiring repeated approval. The issue of consent in research has been a topic of significant discussion in recent times, following the introduction of General Data Protection Regulation (GDPR) in 2018. This was closely followed in Ireland by the supplementary Health Research Regulations, which required GDPR explicit consent for usage of identifiable personal data in health research, a requirement that has been highlighted as causing significant difficulties in research studies (40, 41).

When asked about the involvement of a commercial entity in the donation process, we found that although the majority of our cohort would not allow this to influence their decision to donate a research sample, there was a sizable proportion for whom this would decrease their willingness. This mirrors other studies, where a negative influence of commercial involvement on donation willingness has previously been documented (32, 33, 42). A survey of men attending urology clinics in Ireland found that although 89% would agree with donating a sample for non-profit research, this was more than halved to 39% when questioned about for-profit research (43).

This is likely related to different levels of trust associated with non-profit and for-profit groups (44), as trust levels have been shown to be key drivers of people’s willingness to donate to research. We observed high levels of reported trust in both the respondent’s healthcare team and the scientific community in our survey, as well as majority agreement that these trust levels are important in influencing their decision to donate. Trust levels in both healthcare and scientific teams have also previously been shown to be positively correlated with an individual’s willingness to donate (27, 30, 31, 36).

A better understanding of trust levels and the ways in which it interacts with an individual’s willingness to participate in research could be a valuable tool in enhancing public support for such initiatives. An analysis of the ‘Your DNA, Your Say’ online survey, which included responses from 36,268 individuals across 22 countries, looked at different measures to demonstrate trustworthiness of organisations engaged in collecting genomic data (45). The authors found that transparency around who would benefit from the research was endorsed by 50% of participants, followed by allowing for withdrawal of data and transparency of who would be able to use the data and for what purpose. Other potential measures such as regulating commercial access of research samples through a respected public institution may also alleviate (although not eliminate) some concerns regarding commercial involvement (46).

The issue of transparency is also touched on in our survey, where the vast majority of participants wished to be made aware of any study findings following their donation, even if this finding is clinically non-actionable. A comprehensive literature review on the topic found the majority of included studies were supportive of the return of findings to participants in genomic research (47). This was broadly consistent across different settings (clinical vs. biobank research), different nature of study findings (study specific vs. incidental) and amongst different stakeholder groups (study participants, general public, clinicians and researchers) although support was generally lower where the findings were clinically non-actionable. Researchers and clinicians also tended to be more conflicted and expressed concerns about the potential impact to those receiving these findings (47).

A limitation of our study was that our cohort was markedly skewed towards female participation. Other studies in this area have observed a similar pattern of higher female participation in surveys (5, 22, 23, 48). Much of our online recruitment was via mailing lists from cancer charities and support groups, which considering the generally observed trend of females being more likely to seek out health information (49, 50), are likely to have a higher female representation. We also did not collect data on where respondents received their treatment. It is possible that those treated in establishments with a greater focus on research (e.g. hospitals with active biobanks) or the presence of support staff such as biobank managers or research nurses, could also have an impact on the opinions and knowledge expressed by our survey respondents.

This study adds to the small pool of existing research on sample donation from people with cancer in an Irish context, confirming previous findings of a high degree of willingness to donate a sample to research as well as the impact of factors such as educational levels, trust and commercialisation (22–24). We did observe some differences between our cohort and other studies within Ireland, such as a greater desire for disclosure of genetic findings and a higher (although not universal) level of acceptance of commercial involvement in the process. This could be explained by variations in the makeup of each cohort. For instance, while previous Irish studies have mostly surveyed people with a current cancer diagnosis within a clinical setting, our study included both people with a current cancer diagnosis and those in remission.

In summary, our results show that people living with cancer in Ireland are extremely receptive to donating biological samples to research, broadly matching results seen from studies in other countries. Similarly, their expressed motivations to donate, valuing of trust in healthcare and research teams, and views on potentially controversial issues such as commercialisation, return of results and consent are in keeping with previous findings. This willingness should not be taken for granted as it is highly dependent on the continual trust and good will between an individual and their healthcare team. Further work around patient and public involvement, as has been called for by several national groups, will hopefully provide increased public awareness around the importance of genomic research and patient participation, ultimately bringing direct benefits to people living with cancer.

## Data Availability

All data produced in the present study are available upon reasonable request to the authors

## Declarations

### Author Contributions

WK, EMK & JC conceptualized the study. All authors gave critical insights and strategic direction to the initiative. SOG & AD designed the survey, with input from JCR, EMK, DM & WK. SOG performed data collection, data analysis and drafted the manuscript. All authors read, provided feedback, and approved the final manuscript.

## Supplementary data

**Figure S1.**
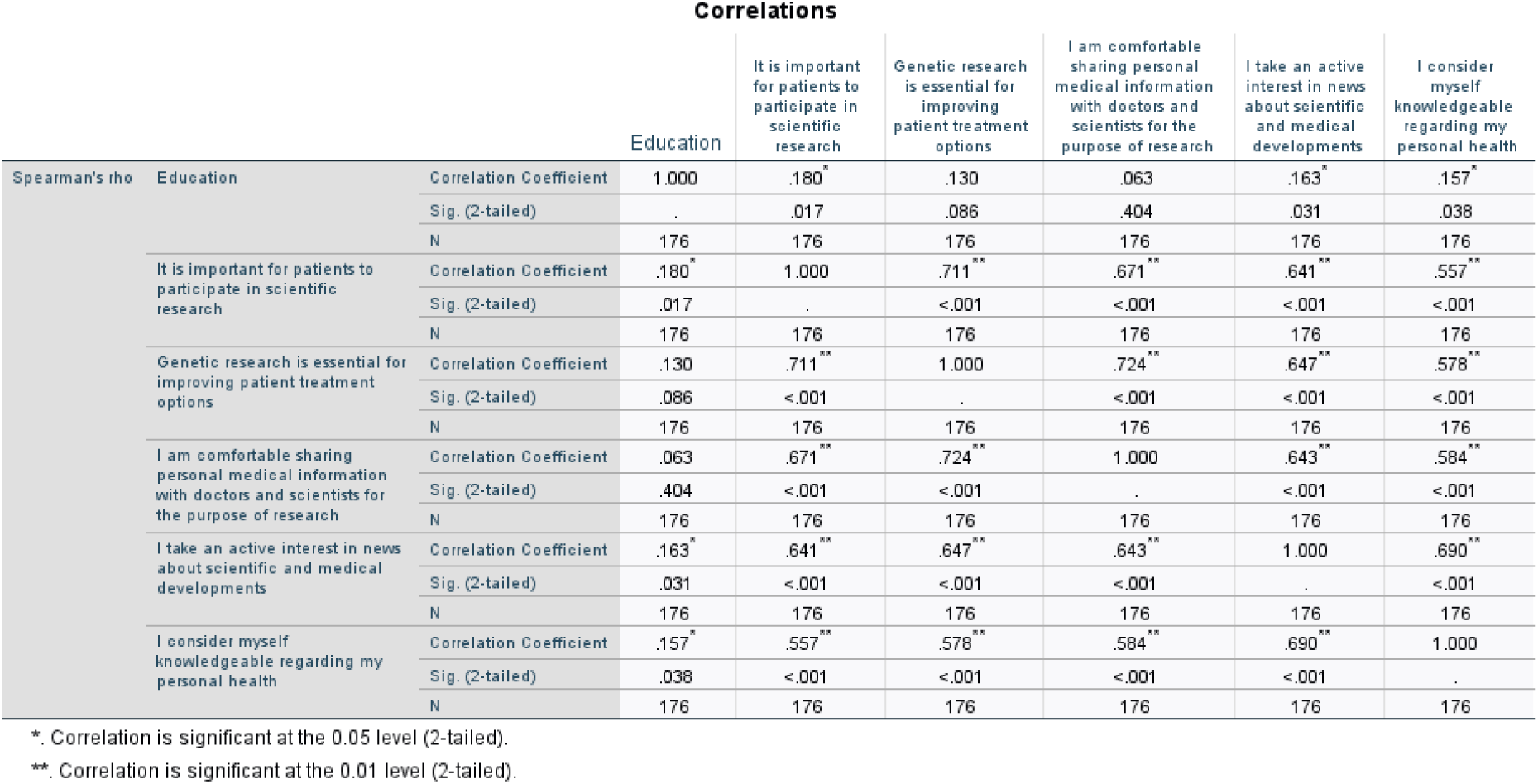
Spearman correlation analysis of educational attainment levels and respondent’s views on research & personal health. Statistical significance is indicated as follows: *p < 0.05, **p < 0.01

